# Determinants of adherence to COVID-19 preventive behaviours in Canada: Results from the iCARE Study

**DOI:** 10.1101/2021.06.09.21258634

**Authors:** Kim L. Lavoie, Vincent Gosselin-Boucher, Jovana Stojanovic, Brigitte Voisard, Geneviève Szczepanik, Jacqueline A. Boyle, Ariane Belanger-Gravel, Simon L. Bacon, for the iCARE Study Team

## Abstract

**Objective:** Key to slowing the spread of SARS-Cov-2 is adherence to preventive behaviours promoted through government policies, which may be influenced by policy awareness, attitudes and concerns about the virus and its impacts. This study assessed determinants of adherence to major coronavirus preventive behaviours, including demographics, attitudes and concerns, among Canadians during the first pandemic wave.

**Methods:** As part of the iCARE study (www.iCAREstudy.com), we weighted data from two population-based, online surveys (April and June, 2020) of Canadian adults. Questions tapped into behaviour change constructs. Multivariate regression models identified determinants of adherence.

**Results:** Data from 6,008 respondents (51% female) were weighted for age, sex, and province. Awareness of government policies was high at both time points (80-99%), and adherence to prevention behaviours was high in April (87.5%-93.5%) but decreased over time, particularly for avoiding social gatherings (68.1%). Adherence was worse among men, those aged 25 and under, and those currently working. Aligned with the Health Beliefs Model, perceptions of the importance of prevention behaviours and the nature of people’s COVID-19-related concerns were highly predictive of adherence. Interestingly, health and social/economic concerns predicted *better* adherence, but having greater personal financial concerns predicted *worse* adherence at both time points.

**Conclusion:** Adherence to COVID-19 prevention behaviours was worse among men, younger adults, and workers, and deteriorated over time. Perceived importance of prevention behaviours measures and health and social/economic concerns predicted better adherence, but personal financial concerns predicted worse adherence. Results have implications for tailoring policy and communication strategies during subsequent pandemic waves.

## Introduction

While we await the widespread dissemination of vaccines, the key to slowing the spread of COVID-19 continues to be maintaining public adherence to the various behaviour-based government prevention measures, e.g., hand washing, physical distancing, avoiding social gatherings, mask wearing, self-isolation if symptomatic or COVID-19 positive, and quarantine after travel.^1,2^ However, adhering to government policies involves making significant personal, social, and economic sacrifices which may undermine people’s motivation to engage in these behaviours.^3^ For example, evidence indicates that adherence to policies that may come with high personal costs (e.g., social/physical distancing) has been less likely (54%) than for other ‘less costly’ preventive behaviours like hand washing (90%).^4^

The extent to which people perceive government policies as important as well as the extent to which people are concerned about the negative consequences of COVID-19, may influence individual decisions to adhere to promoted behaviours. These factors can be summarised by two related but complementary behaviour prediction models: (1) *The Capability, Opportunity, Motivation-Behaviour (COM-B) Model*,^2,5^ which predicts that behaviour change depends on awareness of prevention measures (capability), the belief that measures are personally relevant and important (motivation), and having the resources required to adopt the behaviour (opportunity); and (2) *The Health Beliefs Model*,^6,7^ which posits that in the context of adopting disease prevention measures, a person’s belief in the personal threat(s) posed by an illness or disease, together with a person’s belief in the importance and effectiveness of recommended behaviours, will predict the likelihood a person will (or will not) adopt a particular behaviour.

During this unprecedented health, social, and economic crisis, the global need for public adherence to behaviour-based government policies remains critical to curb the spread of the SARS-Cov-2 virus. Understanding the public’s awareness of and attitudes towards local prevention policies, their COVID-19-related concerns (which reflect risk perceptions), and how these are associated with adherence to prevention behaviours, can be used to inform effective policy planning and communication. Using data that were weighted for age, sex, and province from two population-based samples of adults who completed cross-sectional online surveys designed to assess constructs presented in the *COM-B* and *Health Beliefs* models, the present study assessed associations between COVID-19 policy attitudes and pandemic-related concerns, and adherence to preventive behaviours among Canadians during the first pandemic wave (April – June 2020) using data from iCARE Study.^8,9^

## Methods

### Participants

The **i**nternational assessment of **<u>C</u>**OVID-19-related **<u>A</u>**ttitudes, concerns **<u>RE</u>**sponses and impacts in relation to public health policies (iCARE) Study is an international, multi-wave, cross-sectional, observational study of public awareness, attitudes, concerns, and responses to public health government policies implemented to reduce the spread of COVID-19 (www.icarestudy.com).^9^ As part of this study, we conducted two population-based online surveys of Canadians aged 18 years and over using a recognized polling firm that recruits participants through their proprietary online panel. This panel includes over 400,000 Canadians, the majority of which (60%) were recruited within the past 10 years. Two thirds of the panel were recruited randomly by telephone, with the remainder recruited via publicity and social media. Using data from Statistics Canada, results were weighted within each province according to the sex and age of the respondents in order to make their profiles representative of the current population within each Canadian province (excluding the 3 territories). Then, the weight of each province was further adjusted to represent their actual weight within the ensemble of the 10 Canadian provinces. Data were collected between April 9^th^ and 20^th^, 2020 (Survey 1) and June 4^th^ and 17^th^, 2020 (Survey 2). This timeframe corresponded to the peak and the end of the 1^st^ pandemic wave in Canada. The iCARE study is led by researchers from the Montreal Behavioural Medicine Centre (MBMC: www.mbmc-cmcm.ca) in collaboration with a team of over 200 international collaborators from more than 40 countries. The primary REB approval was obtained from the Comité d’éthique de recherche du Centre intégré universitaire de santé et de services sociaux du Nord-de-l’île-de-Montréal (CIUSSS-NIM), approval # : 2020-2099 / 25-03-2020.

### Survey design

The surveys included 45 questions (Survey 1) and 57 questions and (Survey 2), respectively, assessing sociodemographic variables, physical/mental health, coronavirus infection status, general health behaviours, awareness of local government and municipal policies (validated by Oxford Policy Tracker information)^10^, perceptions and attitudes about policies, concerns about the virus and its impacts, and behavioural responses (adherence) to local government policies. Questions assessing awareness, attitudes (perceived importance), concerns, and behavioural responses were chosen to align with the constructs in both the COM-B^5^ and Health Belief Models.^6,7^ The survey questions can be found at: www.osf.io/nswcm.

### Statistical analysis

Descriptive statistics (weighted means, SD, and proportions with 95% confidence intervals) were calculated to provide an overview of the study sample in terms of demographic characteristics. Questionnaire items that included an answer ‘*I don’t know/I prefer not to answer/Not applicable’* were recoded as missing values. In order to assess adherence to preventive behaviours and COVID-19-related concerns, we computed proportions of individuals that reported practicing behaviours *‘most of the time’* and expressing concerns *‘to a great extent’*, vs. all other categories (“somewhat/very little/not at all”). The following behaviours were considered: handwashing, social distancing (i.e., staying at least 1-2 meters away from others), avoiding social gatherings, avoiding unnecessary travel, mask wearing, self-isolating if symptomatic or COVID-19 positive, and self-quarantining if returning from travel. T-tests and Pearson χ2 statistics (χ2) were used for the comparison of individuals across different strata.

To cluster COVID-19-related concerns, we performed a principal component analysis (PCA) on a polychoric correlation matrix of the 14 (Survey 1) and 18 (Survey 2) variables in the COVID-19 concerns module (ordinal scale). An orthogonal (varimax) rotation was done in order to distribute the factor loadings. We identified concern patterns based on the Kaiser criterion (eigenvalue>1.0), scree plot, and components interpretability.^11^ Items with loadings higher than 0.4 were used to interpret each component of COVID-19 concerns. We observed a three-component structure that included: ‘Health concerns’, ‘personal financial concerns’ and ‘social and economic concerns’ (see details in **Supplement 1**). Mean values (M) and standard deviations (SD) for each of the three components are reported as a score out of four, from 1= “not at all” to 4 = “to a great extent”. Internal consistency for the entire concerns module was excellent for both Survey 1 (α=0.89) and 2 (α=0.91), and the internal consistency of the three factors was excellent (health concerns α=0.88-0.92), very good (personal financial concerns α=0.83-0.85) and satisfactory (social/economic concerns α=0.66-0.76).

Multivariate logistic regression models were performed to assess associations between COVID-19-related policy attitudes and pandemic concerns, and adherence to major preventive measures adjusting for age, sex, education, current employment and province. We classified province into two levels according to COVID-19 cumulative incidence rates as reported by Health Canada^12^ on April 20^th^ and June 12^th^ (final days of data collection for each survey). Specifically, we used the values of the interquartile range (IQR) of the cumulative incidence rates to classify provinces into provinces with rates above and below the national median case rate. Each variable was considered as a separate independent variable, whereas dependent variables included ***good adherence***, defined as those reporting adherence to all three major public health measures (hand washing, social distancing, and avoiding social gatherings) ‘most of the time’ compared to all other responses (outcome 1), and ***adherence to self-isolation*** if you have or believe you have the virus, defined as those reporting adherence ‘most of the time’ compared to all other responses (outcome 2). All statistical tests were two-sided and a p-value < 0.05 was considered as statistically significant. Statistical analysis was performed in SAS, version 9.4.

## Results

### Participant characteristics

A summary of participant characteristics for Survey 1 and 2 can be found in **table 1** and **figure 1**. A total of 16,349 and 17,722 invitations were sent out for Survey’s 1 and 2, and we received 3003 and 3005 completed surveys, respectively, yielding a cooperation rate of 19% (Survey 1) and 18% (Survey 2). The field report showed that the mean survey duration was 12 minutes. Respondents across both surveys were 51.6% female (range 18-95 years) with a mean age 47.2 [SD 17.3] years. Compared to census data available through Statistics Canada, participants in both surveys were well distributed across provinces, age groups, employment status (pre-pandemic) and income, and we had equal proportions of men and women. However, those with high school or less education and with incomes in the top third were marginally less represented.

**Table 1.** Participant characteristics of Canadian respondents to iCARE study online Survey 1 and Survey 2 (April & June 2020).

| Descriptive characteristics | Survey 1 (N=3003) |  |  | Survey 2 (N=3005) |  |  |
| --- | --- | --- | --- | --- | --- | --- |
|  | Unweighted<br>N (%) | Weighted<br>% | 95% CI | Unweighted<br>N (%) | Weighted<br>% | 95% CI |
| **Sex** |  |  |  |  |  |  |
| *Man* | 1484 (49.5) | 48.4 | 46.6 - 50.3 | 1439 (48.0) | 48.4 | 46.5 - 50.2 |
| *Woman* | 1514 (50.5) | 51.6 | 49.7 - 53.4 | 1555 (51.9) | 51.6 | 49.8 - 53.5 |
| **Age** |  |  |  |  |  |  |
| *Less than or equal to 25 years* | 288 (9.7) | 11.9 | 10.6 - 13.3 | 353 (11.9) | 12.4 | 11.1 - 13.7 |
| *26-50 years* | 1374 (46.1) | 42.1 | 40.3 - 43.9 | 1342 (45.2) | 41.7 | 39.9 - 43.5 |
| *51 years or more* | 1316 (44.2) | 46.0 | 44.1 - 47.8 | 1273 (42.9) | 45.9 | 44.1 - 47.8 |
| **Education level** |  |  |  |  |  |  |
| *High school or lower* | 946 (31.8) | 32.2 | 30.5 - 34.0 | 951 (32.0) | 33.1 | 31.3 - 34.9 |
| *Graduate or Postgraduate degree* | 2030 (68.2) | 67.8 | 66.0 - 69.5 | 2025 (68.0) | 66.9 | 65.1 - 68.7 |
| **Region (Province)** |  |  |  |  |  |  |
| *British Columbia* | 401 (13.3) | 13.6 | 12.3 - 14.8 | 402 (13.4) | 13.6 | 12.3 - 14.8 |
| *Alberta* | 351 (11.7) | 11.2 | 10.1 - 12.3 | 351 (11.7) | 11.2 | 10.1 - 12.3 |
| *Saskatchewan* | 86 (2.9) | 3.0 | 2.3 - 3.7 | 96 (3.2) | 3.0 | 2.4 - 3.6 |
| *Manitoba* | 115 (3.8) | 3.5 | 2.9 - 4.2 | 104 (3.5) | 3.5 | 2.8 - 4.2 |
| *Ontario* | 1160 (38.6) | 38.4 | 36.6 - 40.2 | 1160 (38.6) | 38.4 | 36.6 - 40.2 |
| *Quebec* | 690 (23.0) | 23.5 | 21.9 - 25.1 | 692 (23.0) | 23.5 | 21.9 - 25.0 |
| *New Brunswick* | 71 (2.4) | 2.2 | 1.6 - 2.7 | 57 (1.9) | 2.2 | 1.6 - 2.8 |
| *Nova Scotia* | 70 (2.3) | 2.7 | 2.0 - 3.4 | 86 (2.9) | 2.7 | 2.1 - 3.3 |
| *Prince Edward Island* | 20 (0.7) | 0.4 | 0.2 - 0.6 | 8 (0.3) | 0.4 | 0.1 - 0.7 |
| *Newfoundland* | 39 (1.3) | 1.5 | 1.0 - 2.0 | 49 (1.6) | 1.5 | 1.1 - 2.0 |
| **Residential area** |  |  |  |  |  |  |
| *Rural or Country area* | 10 (19.6) | 19.0 | 7.3 - 30.8 | 9 (13.4) | 14.0 | 5.0 - 23.0 |
| *Suburban or Regional* | 18 (35.3) | 37.5 | 22.8 - 52.1 | 22 (32.8) | 38.9 | 25.4 - 52.5 |
| *Urban or City* | 23 (45.1) | 43.5 | 28.8 - 58.1 | 28 (11.9) | 47.0 | 33.4 - 60.7 |
| **Current employment status** |  |  |  |  |  |  |
| *No* | 1451 (49.1) | 50.3 | 48.5 - 52.2 | 1338 (45.3) | 47.0 | 45.1 - 48.9 |
| *Yes* | 1503 (50.9) | 49.7 | 47.8 - 51.5 | 1617 (54.7) | 53.0 | 51.1 - 54.9 |
| **Employment status prior to the COVID-19 pandemic** |  |  |  |  |  |  |
| *Unemployment* | 1038 (36.1) | 37.67 | 35.8 - 39.5 | 944 (33.0) | 35.6 | 33.7 - 37.4 |
| *Employment* | 1690 (58.8) | 59.2 | 57.3 - 61.1 | 1787 (62.6) | 62.1 | 60.2 - 63.9 |
| *Student* | 83 (2.9) | 3.1 | 2.4 - 3.8 | 72 (2.5) | 2.3 | 1.8 - 2.9 |
| **Perceived average annual household income** |  |  |  |  |  |  |
| *Bottom third* | 782 (28.5) | 28.5 | 26.7 - 30.2 | 704 (25.7) | 25.9 | 24.2 - 27.6 |
| *Middle third* | 1568 (57.1) | 57.3 | 55.4 - 59.2 | 1574 (57.5) | 57.4 | 55.5 - 59.3 |
| *Top third* | 397 (14.4) | 14.2 | 12.9 - 15.6 | 461 (16.8) | 16.7 | 15.2 - 18.1 |
| **Health condition at risk<sup>a</sup>** |  |  |  |  |  |  |
| *No* | 1590 (53.5) | 53.7 | 51.8 - 55.5 | 1783 (59.9) | 59.3 | 57.5 - 61.1 |
| *Yes* | 1384 (46.5) | 46.3 | 44.5 - 48.2 | 1195 (40.1) | 40.7 | 38.9 - 42.5 |
| **Presence of any depressive disorder (e.g., major depression)** |  |  |  |  |  |  |
| *No* | 2433 (83.3) | 83.6 | 82.2 - 85.0 | 2532 (86.4) | 86.7 | 85.5 - 88.0 |
| *Yes* | 486 (16.6) | 16.4 | 15.0 - 17.8 | 398 (13.6) | 13.2 | 12.0 - 14.5 |
| **Presence of any anxiety disorder (e.g., panic disorder, generalized anxiety disorder, PTSD)** |  |  |  |  |  |  |
| *No* | 2297 (78.4) | 78.6 | 77.1 - 80.2 | 2370 (80.9) | 81.3 | 79.8 - 82.7 |
| *Yes* | 633 (21.6) | 21.3 | 19.8 - 22.9 | 559 (19.1) | 18.7 | 17.3 - 20.2 |
| **Essential service worker<sup>b</sup>** |  |  |  |  |  |  |
| *No* | 798 (54.7) | 54.1 | 51.5 - 56.8 | 957 (60.6) | 60.8 | 58.3 - 63.2 |
| *Yes* | 662 (45.3) | 45.9 | 43.2 - 48.5 | 622 (39.4) | 39.2 | 36.7 - 41.7 |
| **Health care worker** |  |  |  |  |  |  |
| *No* | 479 (72.8) | 73.3 | 69.8 - 76.8 | 458(73.7) | 74.7 | 71.1 - 78.2 |
| *Yes* | 179 (27.2) | 26.7 | 23.2 - 30.2 | 163(26.2) | 25.3 | 21.8 - 28.9 |
| **Attempt for SARS-Cov-2 testing** |  |  |  |  |  |  |
| *No* | 2802 (94.0) | 94.1 | 93.3 - 95.0 | 2707 (90.8) | 91.1 | 90.1 - 92.2 |
| *Yes, and I got tested* | 118 (4.0) | 3.8 | 3.1 - 4.6 | 197 (6.6) | 6.3 | 5.4 - 7.1 |
| *Yes, but I did not get tested* | 60 (2.0) | 2.0 | 1.5 - 2.5 | 77 (2.6) | 2.6 | 2.0 - 3.2 |
| **SARS-Cov-2 test results<sup>c</sup>** |  |  |  |  |  |  |
| *Positive* | 30 (26.3) | 27.2 | 18.4 - 36.0 | 11 (5.7) | 5.7 | 2.3 - 9.1 |
| *Negative* | 68 (59.6) | 59.6 | 50.1 - 70.0 | 178 (91.7) | 91.6 | 87.5 - 95.7 |
| *I am still waiting for my result* | 16 (14.0) | 13.2 | 6.8 - 19.6 | 5 (2.6) | 2.7 | 0.2 - 5.1 |

<sup>a</sup>Health condition that increases COVID-19 risk: any heart disease or history of heart attack or stroke, any chronic lung disease (e.g., asthma, chronic obstructive pulmonary disease, emphysema/chronic bronchitis); active/current cancer; hypertension; diabetes; severe obesity; any autoimmune disease (e.g., lupus, multiple sclerosis, rheumatoid arthritis, psoriasis, Crohn's disease, inflammatory bowel disease); <sup>b</sup>responded only by the individuals who are currently working; <sup>c</sup>responded only by the individuals who got tested for COVID-19

**Figure 1.**
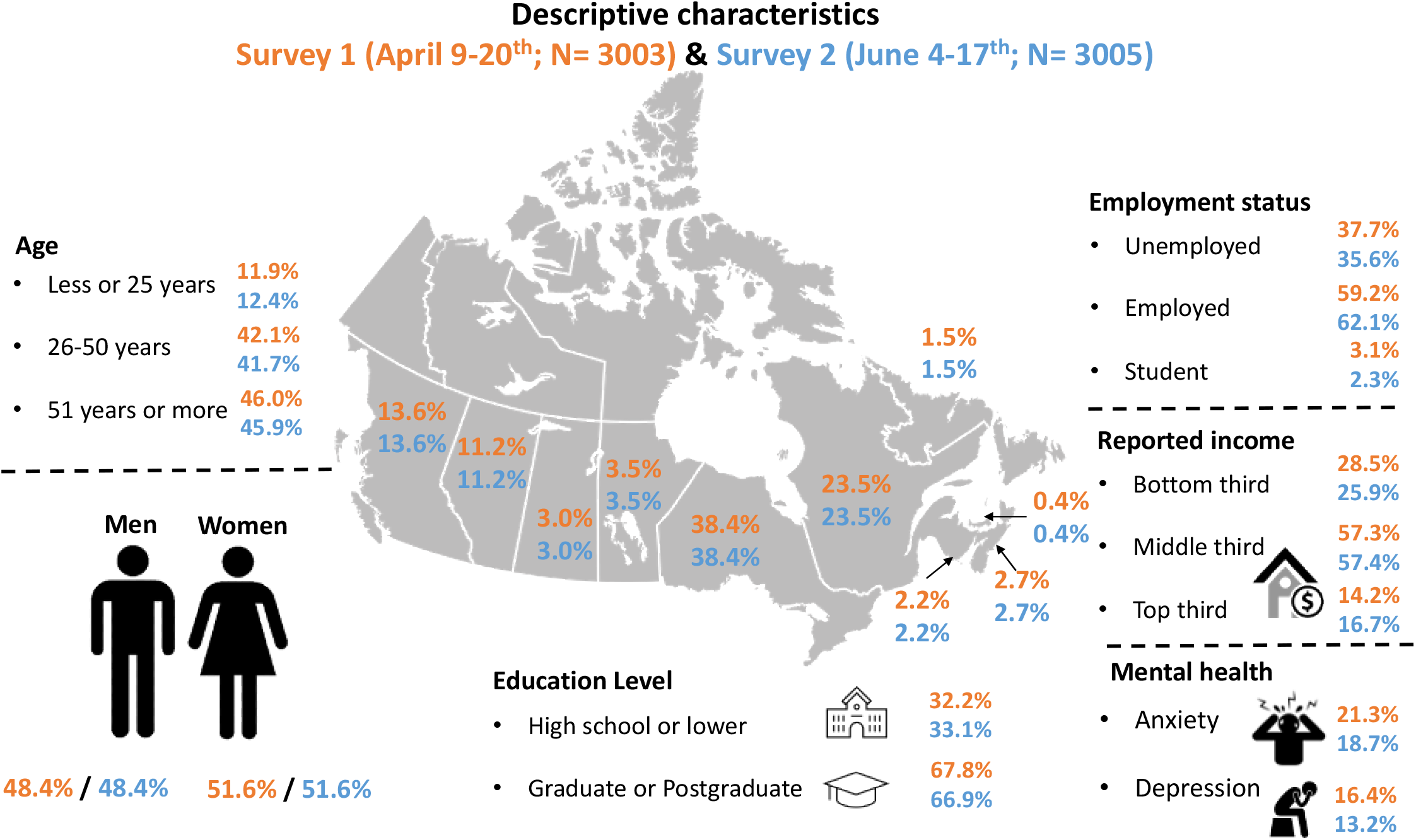
Selected sociodemographic characteristics of 6008 Canadian participants in two online surveys in the iCARE study in April and June 2020.

### Awareness of public health measures

We assessed Canadians’ ***awareness*** of government policies by asking respondents *what actions or behaviours their local government or health authority has recommended* to reduce and slow the spread of COVID-19 (see **Supplement table S1**). Overall, the vast majority of respondents (96.7%-98.1%) were aware of the major recommendations in April (e.g., hand washing, staying 1-2 metres away from others, self-isolation if COVID-19+ or symptomatic). Awareness levels remained high in June (96.5% to 98.4%) for all behaviours except avoiding social gatherings - which decreased to 68.1%.

### Adherence to preventions behaviours

We assessed Canadians’ adherence to the various recommended behaviours by asking respondents *the frequency with which they had engaged in each preventive behaviour in the previous 7 days*. A summary of the proportion of people reporting engaging in the different public health measures at least most of the time in April (Survey 1) and June (Survey 2) is presented in **table 2**. Overall, the majority of respondents reported being adherent to recommended behaviours in April, though this varied by behaviour. At least 87.5-93.5% of Canadians reported hand washing, social distancing, avoiding social gatherings, avoiding non-essential travel, self-isolating if symptomatic or infected by the coronavirus, and self-quarantining if returning from travel most of the time. By June, we observed a general decrease in adherence to most recommended behaviours, including hand washing (−7.7%), social distancing (−8.2%), avoiding non-essential travel (−8.9%), and avoiding social gatherings (−23.7%). The only behaviours that remained relatively stable over time were self-isolating if you have or believe you have the virus (−0.6%), and quarantining after returning from a trip (−0.9). Though mask wearing was not formally recommended in April or June in most provinces, 22.5% of Canadians in April and 67.7% in June reported wearing masks at least most of the time, and was the only preventive behaviour to increase over time. All proportions and 95% confidence intervals are presented in **table 2**.

**Table 2.**
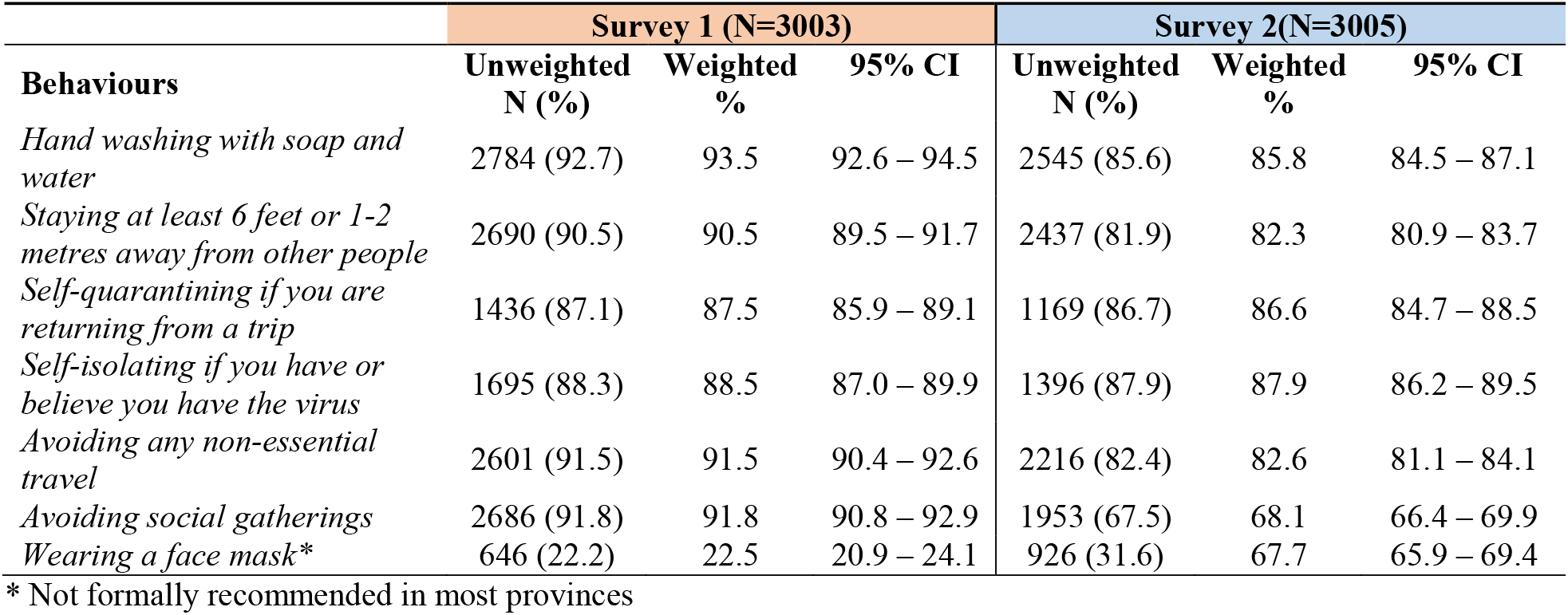
Overall frequency and proportion of Canadian respondents to online surveys in the iCARE study indicating adherence to preventive behaviours ‘Most of the time’ in the past 7 days in April and June 2020.

| **Behaviours** | **Survey 1 (N=3003)** |  |  | **Survey 2(N=3005)** |  |  |
| --- | --- | --- | --- | --- | --- | --- |
|  | **Unweighted**<br>**N (%)** | **Weighted**<br>**%** | **95% CI** | **Unweighted**<br>**N (%)** | **Weighted**<br>**%** | **95% CI** |
| *Hand washing with soap and water* | 2784 (92.7) | 93.5 | 92.6 – 94.5 | 2545 (85.6) | 85.8 | 84.5 – 87.1 |
| *Staying at least 6 feet or 1-2 metres away from other people* | 2690 (90.5) | 90.5 | 89.5 – 91.7 | 2437 (81.9) | 82.3 | 80.9 – 83.7 |
| *Self-quarantining if you are returning from a trip* | 1436 (87.1) | 87.5 | 85.9 – 89.1 | 1169 (86.7) | 86.6 | 84.7 – 88.5 |
| *Self-isolating if you have or believe you have the virus* | 1695 (88.3) | 88.5 | 87.0 – 89.9 | 1396 (87.9) | 87.9 | 86.2 – 89.5 |
| *Avoiding any non-essential travel* | 2601 (91.5) | 91.5 | 90.4 – 92.6 | 2216 (82.4) | 82.6 | 81.1 – 84.1 |
| *Avoiding social gatherings* | 2686 (91.8) | 91.8 | 90.8 – 92.9 | 1953 (67.5) | 68.1 | 66.4 – 69.9 |
| *Wearing a face mask\** | 646 (22.2) | 22.5 | 20.9 – 24.1 | 926 (31.6) | 67.7 | 65.9 – 69.4 |

\* Not formally recommended in most provinces

We assessed adherence to all three major public health measures (i.e., the proportion of people reporting hand washing, social distancing, and avoiding social gatherings at least most of the time), referring to this as ‘***good adherence’***. Using this classification, a total of 83.1% (95% CI 81.8-84.7) of Canadians reported good adherence in April, but this decreased significantly to 55.7% (95% CI 54.6-58.3) by June. As detailed in **figure 2a**, fully adjusted analyses revealed men (compared to women) were 50% less likely to have good adherence compared to women in April (OR_adj_ 0.50, 95% CI 0.41-0.61) and 22% less likely in June (OR_adj_ 0.78, 95% CI 0.67-0.91). Further, compared to those aged 51 years and over, those aged 25 and younger and aged 25 to 50 years were 65% (OR_adj_ 0.35, 95% CI 0.26-0.49) and 46% (OR_adj_ 0.54, 95% CI 0.42-0.68) less likely to have good adherence respectively, in April and 70% (OR_adj_ 0.30 95% CI 0.23-0.39) and 42% (OR_adj_ 0.58, 95% CI 0.49-0.70) less likely to have good adherence respectively, in June. Compared to those not currently working, those currently working were 26% less likely to have good adherence in April (OR_adj_ 0.74, 95% CI 0.59-0.92) but not June (OR_adj_ 0.91, 95% CI 0.77-1.08). Finally, those living in provinces above the median number of national cases were 38% less likely to have good adherence in April (OR_adj_ 0.62, 95% CI 0.0.39-0.99) but not June (OR_adj_ 1.08, 95% CI 0.80-1.45). There was no effect for education.

**Figure 2a.**
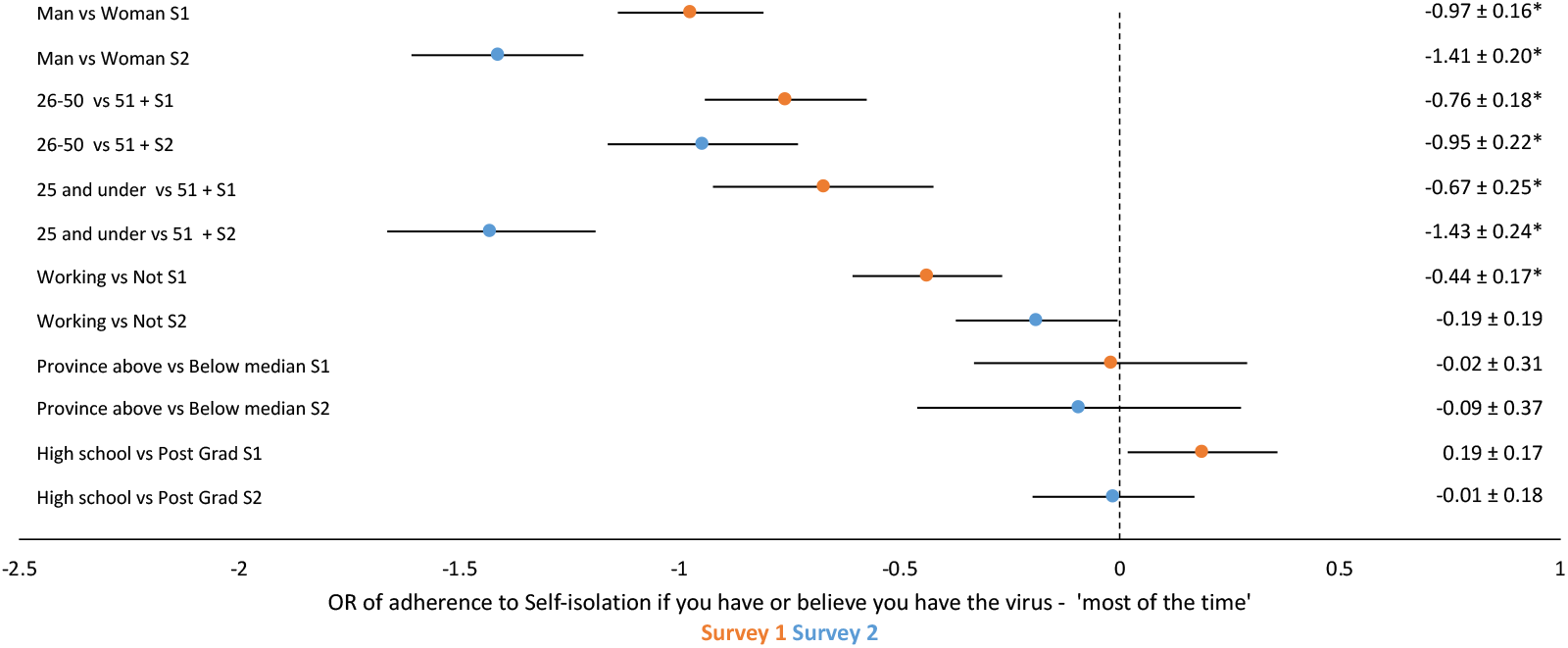
Forest plot of adjusted odd ratio of adherence to *preventive behaviours* ‘Most of the time’ as a function of sociodemographic characteristics among Canadian participants in two online surveys in the iCARE study in April and June 2020.

We repeated these analyses examining adherence to self-isolation if symptomatic or infected and found that 88.5% (95% CI 87.0-89.9) and 87.9% (95% CI 86.2-89.5) reported doing this ‘most of the time’ in April and June, respectively. We also repeated the fully adjusted analysis as a function of the same demographics and observed the same general pattern of results: in both April and June, men (April: OR_adj_ 0.38, 95% CI 0.0.67-0.91; June: OR_adj_ 0.24, 95% CI 0.24-0.36) and those aged 25 and younger (April: OR_adj_ 0.51, 95% CI 0.31-0.84; June OR_adj_ 0.24, 95% CI 0.15-0.38) were less likely to self-isolate compared to women and those aged 25 and older. Further, those currently working in April (OR_adj_ 0.65, 95% CI 0.46-0.90), but not June (OR_adj_ 0.83, 95% CI 0.58-1.19), were less likely to self-isolate compared to those not working (see **figure 2b**).

**Figure 2b.**
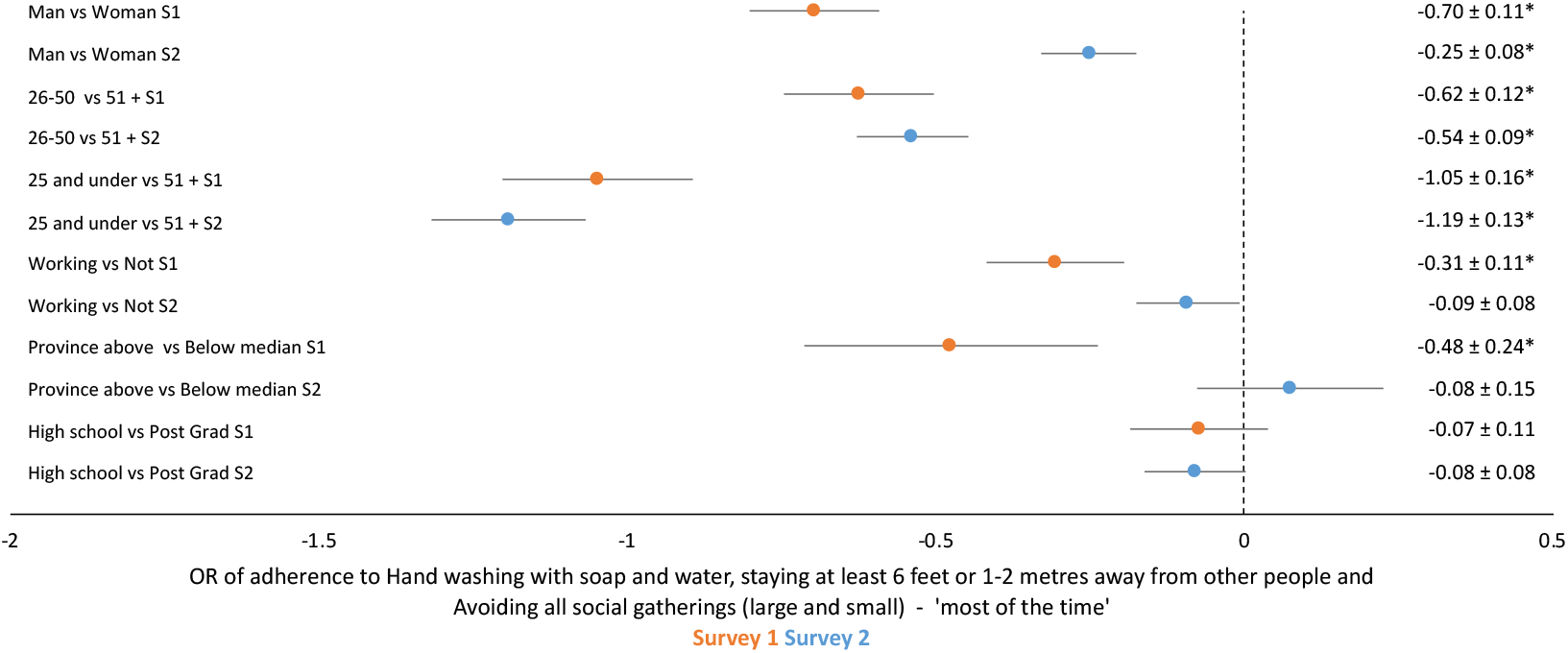
Forest plot of adjusted odd ratio of adherence to *self-isolation* ‘Most of the time’ time’ as a function of sociodemographic among Canadian participants in two online surveys in the iCARE study in April and June 2020. * Defined as: Hand washing with soap and water and staying at least 6 feet or 1-2 metres away from other people and avoiding social gatherings; *CI*: Confidence Interval; *OR*: Odds Ratio; Adjusted for: age, sex, education, current employment, province.

### Perceived importance of recommended behaviours and associations with adherence

We assessed Canadians’ perceptions of the ***importance*** of recommended behaviours by asking respondents to *what extent they believe that the measures asked of them by their government or local health authority are important* for preventing or reducing the spread of the SARS-Cov-2 virus. Though the majority (i.e., 87%, 95% CI 86.2-88.6) perceived recommended behaviours to be very important in April, this diminished significantly by June (72%, 95% CI 70.8-74.1). Further, men and those aged 25 years or younger perceived recommended behaviours to be significantly less important than women and those over aged 25 at both time points (**figure 3**). As detailed in **table 3**, multivariate analyses adjusting for age, sex, education level, current employment status, and province revealed a 4.0-fold (OR_adj_ 4.01 95% CI 3.12-5.14) and 3.3-fold (OR_adj_ 3.34 95% CI 2.79-3.98) increased odds of good adherence ‘most of the time’ among those who perceived recommended behaviours to be ‘extremely important’ in April and June, respectively. The same pattern was observed for self-isolation: a 3-fold (OR_adj_ 3.38 95% CI 2.39-4.78) and 2.8-fold (OR_adj_ 2.85 95% CI 2.04-3.99) increased odds of self-isolating ‘most of the time’ was seen among those who perceived recommended behaviours to be ‘extremely important’ in April and June, respectively.

**Figure 3.**
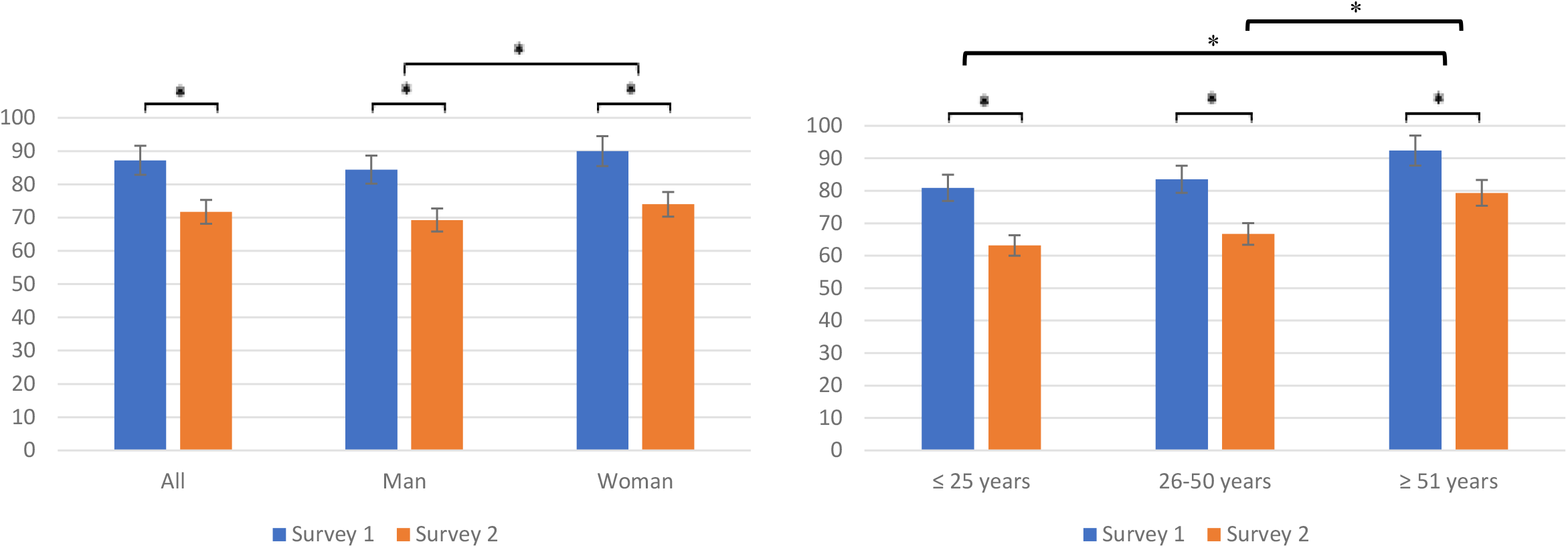
Proportions (95% CIs) perceiving public health measures ‘very important’ as a function of sex and age among Canadian participants to two online surveys in the iCARE study in April and June 2020. All proportions significantly different from one another except for ≤25 years compared to 25-50 years

**Table 3.** Adjusted odd ratios for adherence to preventive behaviours ‘most of the time’ as a function of high perceived importance of government prevention measures among Canadian respondents to online surveys in the iCARE study in April and June 2020.

|  | Survey 1(N=3003) |  |  | Survey 2(N=3005) |  |  | Survey 1(N=3003) |  |  | Survey 2 (N=3005) |  |  |
| --- | --- | --- | --- | --- | --- | --- | --- | --- | --- | --- | --- | --- |
| Behaviour | Composite: Good adherence to major prevention behaviours* |  |  |  |  |  | Self-isolating if you have or believe you have the virus |  |  |  |  |  |
| Variable | OR | 95% CIs |  | OR | 95% CIs |  | OR | 95% CIs |  | OR | 95% CIs |  |
| High perceived importance of government prevention measures | 4.01 | 3.13 | 5.14 | 3.34 | 2.79 | 3.98 | 3.38 | 2.39 | 4.78 | 2.85 | 2.04 | 3.99 |

\*Defined as: Hand washing with soap and water and staying at least 6 feet or 1-2 metres away from other people and avoiding social gatherings; CI: Confidence Interval; OR: Odds Ratio; Adjusted for: age, sex, education, current employment, province.

### COVID-19-related concerns and associations with adherence

We assessed the nature and extent of Canadians’ COVID-19-related concerns. A summary of the proportion of people reporting having each concern “to a great extent” can be found in **Supplement table S2**. As detailed in **table 4**, our PCA analyses revealed that after adjustment for age, sex, education, current employment status, and province, the concern type with the highest mean score (out of 4) in April was social/economic concerns (M±SD =3.12±0.65), followed closely by health concerns (M±SD=3.02±0.73) and personal financial concerns (M±SD=2.62±0.88). By June, concern levels across all three types had decreased significantly, though the absolute magnitude of these decreases was nominal. Women had significantly *higher* levels of concerns across all types, and those in the top third of annual household income had *lower* levels of health and personal financial concerns across both time points (p’s<.01). Those aged 51 years and above had significantly *higher* levels of health and significantly *lower* levels of personal financial concerns across both time points (p’s<.01). Those with a graduate/postgraduate degree had significantly *higher* levels of social/economic concerns at both time points, and had significantly *higher* health and personal financial concerns by June (p’s<.05). Finally, those who were employed had *higher* levels of all concerns types at both time points, with the exception of greater personal financial concerns in April. All means and variance measures are presented in **table 4**.

**Table 4.**
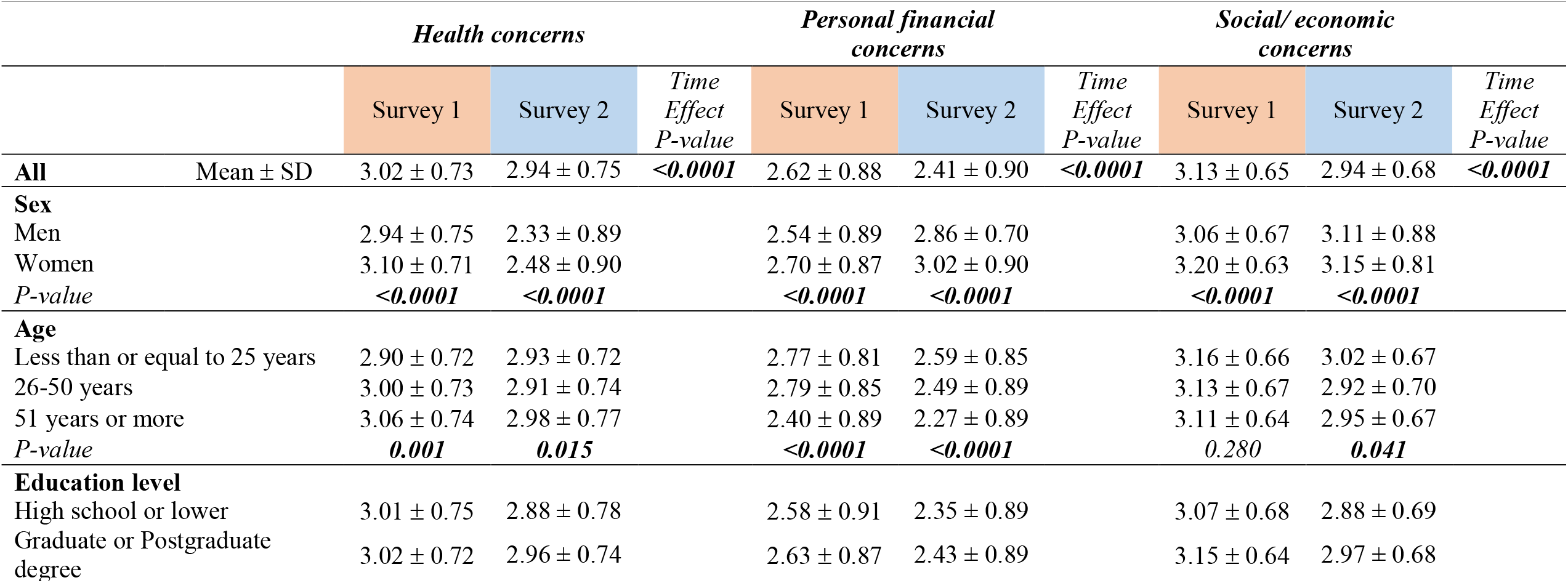
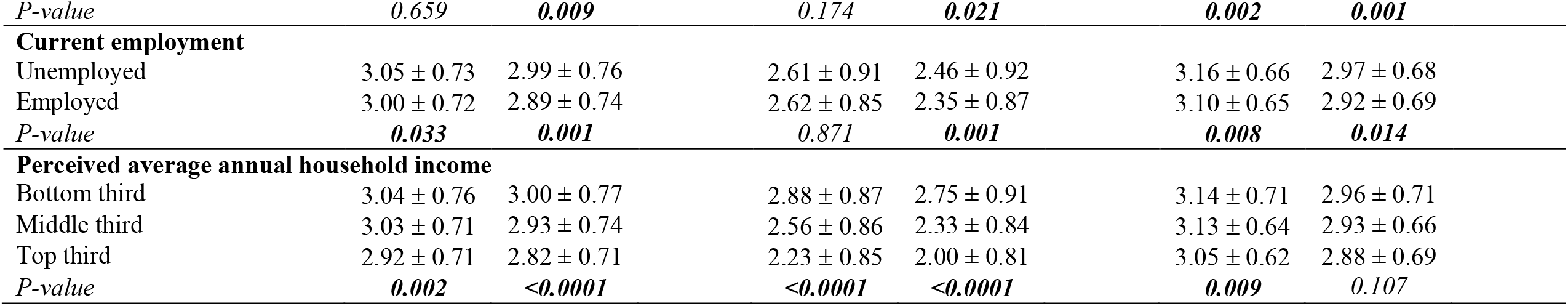
Means and standard deviations for COVID-19-related concerns of as function of sociodemographic characteristic among Canadian respondents to online surveys in the iCARE study in April and June 2020.

We assessed the association between **concern type** and ‘good adherence’ to the three major preventive behaviours in multivariate analyses adjusting for age, sex, education level, current employment status, and province. As detailed in **table 5**, results revealed that both high health concerns (OR_adj_ 1.81 95% CI 1.53-2.12) and high social-economic concerns (OR_adj_ 2.15 95% CI 1.89-2.46) were associated with significantly better adherence to the major prevention measures in April, but only high health concerns (OR_adj_ 2.05 95% CI 1.60-2.61) was associated with better adherence in June. Interestingly, high personal financial concerns (OR_adj_0.82 95% CI 0.71-0.95) were associated with significantly *worse adherence* in April, but this was no longer significant by June (OR_adj_ 0.92 95% CI 0.83-1.03). A similar and even more consistent pattern was seen for adherence to ‘self-isolation’ if symptomatic or coronavirus+. At both time points, both high health (April: OR_adj_2.05 95% CI 1.60-2.61; June: OR_adj_ 2.46 95% CI 1.86-3.25) and high social/economic concerns (April: OR_adj_ 1.35 95% CI 1.05-1.75; June: OR_adj_ 1.37 95% CI 1.03-1.85) were associated with significantly greater adherence to self-isolation, and having high personal financial concerns (April: OR_adj_ 0.78 95% CI 0.63-0.98; June: OR_adj_ 0.61 95% CI 0.48-0.79) was associated with significantly *worse adherence* to self-isolation.

**Table 5.**
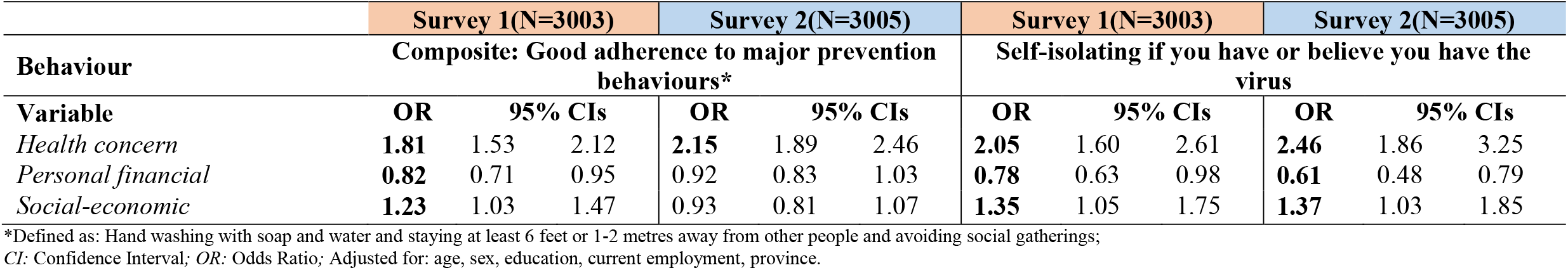
Adjusted odd ratios for adherence to preventive behaviours ‘most of the time’ as a function of different sets of concern among Canadian respondents to online surveys in the iCARE study in April and June 2020.

|  | Survey 1(N=3003) |  |  | Survey 2(N=3005) |  |  | Survey 1(N=3003) |  |  | Survey 2(N=3005) |  |  |
| --- | --- | --- | --- | --- | --- | --- | --- | --- | --- | --- | --- | --- |
| Behaviour | Composite: Good adherence to major prevention behaviours* |  |  |  |  |  | Self-isolating if you have or believe you have the virus |  |  |  |  |  |
| Variable | OR | 95% CIs |  | OR | 95% CIs |  | OR | 95% CIs |  | OR | 95% CIs |  |
| Health concern | 1.81 | 1.53 | 2.12 | 2.15 | 1.89 | 2.46 | 2.05 | 1.60 | 2.61 | 2.46 | 1.86 | 3.25 |
| Personal financial | 0.82 | 0.71 | 0.95 | 0.92 | 0.83 | 1.03 | 0.78 | 0.63 | 0.98 | 0.61 | 0.48 | 0.79 |
| Social-economic | 1.23 | 1.03 | 1.47 | 0.93 | 0.81 | 1.07 | 1.35 | 1.05 | 1.75 | 1.37 | 1.03 | 1.85 |

\*Defined as: Hand washing with soap and water and staying at least 6 feet or 1-2 metres away from other people and avoiding social gatherings; *CI*: Confidence Interval; *OR*: Odds Ratio; Adjusted for: age, sex, education, current employment, province.

## Discussion

We identified determinants of adherence to major SARS-CoV-2 preventive behaviours, including demographics, attitudes, and concerns, among Canadians during the first pandemic wave (April – June 2020). Results indicated high awareness of prevention behaviours at both time points. Adherence to prevention behaviours was also high at both time points *except* for avoiding social gatherings, which dropped from 91.8% in April to 67.9% in June. The fact that this sharp decline was observed before most provinces relaxed prevention measures indicates that policies that require making important social sacrifices, like avoiding social interaction with family and friends, may be particularly difficult for people and populations to sustain over time. This hypothesis is supported by previous research^4^ as well as iCARE data from Australia,^13^ which also showed steep declines in avoidance of social gatherings between May (81%) and July (48%) relative to other, less personally costly measures like hand washing (May = 78%; July = 72%).^13^ Supporting this hypothesis is the sharp increases in mask wearing (the only prevention behaviour to increase over time) observed between April and June, which went from 22.5% to 67.7%. This is impressive considering that mask wearing was not formally recommended in April or June in most provinces,^10^ and suggests that measures that either don’t involve making important social sacrifices, or those that may enable people to gather more safely, might be more readily adopted. Consistent with the COM-B model^2,5^, masks were also more readily available by June, which possibly enhanced people’s opportunity and capacity to engage in the behaviour.

Interestingly, our data revealed that adherence to major preventive behaviours, including self-isolation if symptomatic or coronavirus+, was significantly worse among men compared to women. This is consistent with other COVID-19 studies,^14,15^ prior pandemic studies,^16^ and iCARE data from Australia.^13,17^ There is also in line with a growing literature demonstrating sex-specific differences not only in the epidemiology of COVID-19,^18,19^ but also in behavioural responses and impacts of the pandemic.^20^ Women have also been consistently shown to demonstrate better adherence to preventive behaviours compared to men.^20-23^ Of note, women in our study also perceived prevention measures to more important relative to men, and reported having significantly higher COVID-19-related concerns across all concern types (health, personal financial, and social/economic). Consistent with the Health Beliefs Model, women may have been more adherent due to their heightened disease risk perception and beliefs about the importance of engaging in prevention measures, which are important determinants of behaviour and are typically lower among men.^24,25^ The higher COVID-19 concerns observed among women may also be ascribed to gender roles, which traditionally have women assuming greater caregiving roles both within and outside the family.^26^ Women are also disproportionately represented in frontline healthcare occupations (e.g., nursing), which places them at increased risk of contracting the virus and transmitting it to others under both their formal and informal care.^27,28^ Finally, women have been shown to suffer greater social, economic, and mental health impacts as a result of the pandemic,^26^ so their higher level of concerns may reflect accurate risk perceptions and their desire to avoid experiencing these negative outcomes.

Poorer adherence to prevention measures, including self-isolation, was also more likely to occur among those age 25 years and under at both time points. Younger age groups (i.e., those under age 30 years) have been consistently shown to demonstrate lower adherence to prevention behaviours compared to older age groups in similar COVID-19 studies,^13-15,17^ possibly due to reports that younger people are at much lower risk of severe disease, hospitalizations, and death.^29-31^ The fact that younger people do seem to experience milder disease and far fewer complications may indeed undermine their motivation to adhere to preventive behaviours, many of which may come at a high cost that may seem to exceed personal benefits. This hypothesis is supported by our findings showing that younger people perceived prevention behaviours to be less important, and had comparatively lower health concerns relative to older age groups. In contrast, younger people were most concerned about the social/economic impacts of the virus. This suggests that policy communication promoting adherence to preventive behaviours targeting young people may need to focus on the positive social and economic outcomes associated with adherence (e.g., greater adherence will reduce transmission and case rates, leading to relaxation of confinement measures and a re-opening of the economy) rather than those focusing on health outcomes that they may find less persuasive.

Poorer adherence to prevention measures, including self-isolation, was also more likely to occur among those currently working in April, but not June. This suggests that those who were still employed at the peak of the first wave may have faced additional barriers to comply with preventive behaviours due to their employment status. We explored the extent to which non-adherence to self-isolation recommendations occurred more frequently among essential service workers, but found no evidence of this at either time point, with both essential and non-essential workers complying roughly 86% of the time (data not shown). Alternatively, it is possible that those who were fortunate enough to have a job during the peak of the first wave may have felt more pressure to work, perhaps due to fears of the negative repercussions of missing work, including job loss and lost revenue.^32^ In support of this hypothesis was our finding linking higher personal financial concerns with *less* adherence to both major preventive behaviours and self-isolation at both time points. This suggests that poor adherence to self-isolation might represent a capability rather than a motivational problem as defined by the COM-B model, where individuals may have wanted to comply with self-isolation recommendations, but felt unable to due to fears of financial losses and/or negative repercussions by employers. Enabling Canadians to overcome financial barriers to adherence to self-isolation by implementing short-term emergency response benefits (e.g., Canada Recovery Sickness Benefit, CRSB) during the isolation period, as well as ensuring that employers will not penalize them with redundancy or other punitive measures may enhance adherence to these behaviours. However, the level of benefits and protection would need to be sufficient. These kinds of measures are also supported by research from other countries (e.g., Israel) indicating that financial compensation can significantly improve adherence to recommended COVID-19 isolation directives.^33^

Of note, how important individuals perceived preventive behaviours to be for reducing the spread of the virus was associated with a 3 to 4-fold increased odds of adherence after adjustment for covariates. This was consistent with iCARE data from Australia, which found that 91% of respondents who reported high adherence to major prevention measures believed them to be ‘very important’, compared to only 22% of those with low adherence.^17^ This finding was also observed in iCARE data from Ireland, which found those who believed prevention measures to be ‘very important’ for reducing virus transmission were also those who exhibited the highest adherence to all prevention behaviours.^34^ These findings are all in line with the Health Beliefs Model,^6,7^ which posits that beliefs about the importance of disease prevention measures are powerful motivators of behaviours, and indicates that adherence levels may be particularly vulnerable to information that suggests prevention behaviours may lack efficacy for reducing virus transmission. This hypothesis is supported by iCARE global data obtained during the first wave (March 27-April 15, n=20,537), which showed that ‘demonstrating how adherence to prevention behaviours are helping to reduce the spread of virus’ emerged as one of the strongest motivators of adherence.^35^ This has implications for policy communication and emphasizes the importance of providing the public feedback on the real value of engaging in preventive behaviours.

Despite Canadians being most concerned about the social/economic impacts of the pandemic, the concern most strongly related to adherence to major preventive behaviours and self-isolation at both time points were health concerns, which were associated with a nearly 2 to 2.5-fold increased likelihood of adherence independent of age, sex, education, employment status, and province. This is consistent with the Health Beliefs Model,^6,7^ which posits that higher concerns about the health consequences of a disease are powerful motivators of behaviours. The health concerns variable is a composite of concerns about personally becoming infected and infection among friends, family, and the community, and suggests that for those not concerned about negative health consequences of the virus, motivating adherence to prevention measures using health-focused messaging may be a tough sell.^36^ Indeed, most of the policy messaging in Canada (and elsewhere) has focused on preventing the negative health consequences of getting COVID-19 (i.e., “you could get sick or worse”) rather than the multitude of personal and collective gains associated with staying healthy, which include the resumption of social activities, better job prospects, and a stronger economy.^37^ As mentioned above, this may not have resonated among younger people in particular, who have likely been exposed to reports indicating that severe illness is much less probably among those in their age group.^29-31^ Repeated exposure to messaging focused on avoidance of negative outcomes, rather than on the achievement of positive outcomes, has also been shown to reduce the efficacy of the message over time.^38^ In fact, more positive messaging has been shown to reinforce good adherence behaviour,^38,39^ by communicating that the measures you are taking are keeping you and your loved ones safe, but also increasing job opportunities, strengthening the economy, and will allow us to resume normal social activities sooner. The effects of positive, rather than negative messaging, has been seen with other complex behaviours like physical activity, ^40^ and may be important for informing policy communication during subsequent pandemic waves.

### Limitations and strengths

This study should be interpreted in light of some methodological limitations. First, although we analysed and weighted large, representative samples of Canadians with representation across age, sex, and province, the absolute number of participants in certain provinces (e.g., Atlantic) was lower, making some inter-provincial comparisons unadvisable. Second, the survey was only available in English and French, which may have led to an underrepresentation of certain non-native English or French speaking groups. Further, our survey included a higher proportion of university-educated individuals, though relatively few assessed themselves to be in the top income tercile. As such, results might not generalize as well to less educated or those from the highest income group. Third, since the survey was voluntary, and participants were drawn from a polling firm’s subject pool, participation may have been subject to some degree of selection bias, though applying the National weightings accounts for some of this, it can never fully remove bias. Finally, data were self-reported, which may have been subject to social desirability bias,^41^ though this may have been mitigated by the fact that the survey was completely anonymous and did not collect any personal identifiable information. Despite some limitations, this study also had a number of important strengths. The survey was designed based on established theories of behaviour change (Health Beliefs Model and COM-B), which is important in the context of identifying targets for intervention and policy implementation. The study collected data during peak lockdown (April) through to early deconfinement (June) in Canada, which allowed for the assessment of changes over time across two critical periods of first wave of the pandemic in Canada. Finally, we used robust statistical methods to determine the factor structure of our variables measuring concerns, which was found to have excellent internal consistency, which is important for ensuring the validity of our results linking concern types to behavioural adherence.

## Conclusions

Awareness of and adherence to major COVID-19 prevention behaviours was high during the first pandemic wave, was worse among men and younger adults, and generally deteriorated over time. Perceived importance of prevention measures decreased over time, but was associated with 3-4-fold increased odd of adherence at both time points. While both health and social/economic concerns predicted *better* adherence to prevention behaviours, having greater personal financial concerns predicted *worse* adherence to prevention behaviours. Results suggest that policy communication promoting adherence to preventive behaviours may need to emphasize the positive health *and* social/economic outcomes associated with adherence, as well as provide feedback on the efficacy of engaging in preventive behaviours for reducing virus transmission to prevent reductions in adherence over time.

## Data Availability

Data from the iCARE Study is open-access at the following link: https://osf.io/nswcm/

## Acknowledgements

The primary source of funding for the iCARE study has been primarily through re-directed funding associated with Montreal Behavioural Medicine Centre, including funds from the Canada Research Chairs Program (950-232522, Chair holder: Dr. Kim L. Lavoie), a Canadian Institutes of Health Research-Strategy for Patient Oriented Research Mentoring Chair (SMC-151518, PI: Dr. Simon L. Bacon), a Fonds de Recherche du Québec: Santé Chair (251618, PI: Dr. Simon L. Bacon), and a Fonds de Recherche du Québec: Santé Senior Research Award (34757, PI: Dr. Kim L Lavoie). Addition support has been provided by the Canadian Institutes of Health Research (MS3- 173099, co-PI’s: Simon L. Bacon & Kim L. Lavoie) and the Fonds de Recherche du Québec: Société et Culture (2019-SE1-252541, PI: Dr. Simon L. Bacon). None of the funders were involved in the study design. We also acknowledge the support from our MBMC iCARE Team, particularly administrative support by Mr. Guillaume Lacoste and Katherine Seguin, and analytic support by Ms. Ruth Bruno.

## iCARE Study collaborators

**Lead investigators**: Kim L. Lavoie, PhD, University of Quebec at Montreal (UQAM) and CIUSSS-NIM, CANADA; Simon L. Bacon, PhD, Concordia University and CIUSSS-NIM, CANADA. **Collaborators** (in alphabetical order by country): ABU DHABI: Zahir Vally, PhD, United Arab Emirates University; ARGENTINA: Nora Granana, PhD, Hospital Durand; Analía Verónica Losada, PhD, University of Flores; AUSTRALIA: Jacqueline Boyle, PhD, Monash University; Joanne Enticott, PhD, Monash University; Shajedur Rahman Shawon, PhD, Centre for Big Data Research in Health, UNSW Medicine; Shrinkhala Dawadi, MSc, Monash University; Helena Teede, MD, Monash University; AUSTRIA: Alexandra Kautzky-Willer, MD, Medizinische Universität Wien; BANGLADESH: Arobindu Dash, MS, International University of Business, Agriculture & Technology; BRAZIL: Marilia Estevam Cornelio, PhD, University of Campinas; Marlus Karsten, Universidade do Estado de Santa Catarina - UDESC; Darlan Lauricio Matte, PhD, Universidade do Estado de Santa Catarina - UDESC; Felipe Reichert, PhD, Universidade; CANADA: Ahmed Abou-Setta, PhD, University of Manitoba; Shawn Aaron, PhD, Ottawa Hospital Research Institute; Angela Alberga, PhD, Concordia University; Tracie Barnett, PhD, McGill University; Silvana Barone, MD, Université de Montréal; Ariane Bélanger-Gravel, PhD, Université Laval; Sarah Bernard, PhD, Université Laval; Lisa Maureen Birch, PhD, Université Laval; Susan Bondy, PhD, University of Toronto - Dalla Lana School of Public Health; Linda Booij, PhD, Concordia University; Roxane Borgès Da Silva, PhD, Université de Montréal; Jean Bourbeau, MD, McGill University; Rachel Burns, PhD, Carleton University; Tavis Campbell, PhD, University of Calgary; Linda Carlson, PhD, University of Calgary; Kim Corace, PhD, University of Ottawa; Olivier Drouin, MD, CHU Sainte-Justine/Université de Montréal; Francine Ducharme, MD, Université de Montréal; Mohsen Farhadloo, Concordia University; Carl Falk, PhD, McGill University; Richard Fleet MD, PhD, Université Laval; Michel Fournier, MSc, Direction de la Santé Publique de Montréal; Gary Garber, MD, University of Ottawa/Public Health Ontario; Lise Gauvin, PhD, Université de Montréal; Jennifer Gordon, PhD, University of Regina; Roland Grad, MD, McGill University; Samir Gupta, MD, University of Toronto; Kim Hellemans, PhD, Carleton University; Catherine Herba PhD, UQAM; Heungsun Hwang, PhD, McGill University; Jack Jedwab, PhD, Canadian Institute for Identities and Migration and the Association for Canadian Studies; Keven Joyal-Desmarais, PhD, Concordia University; Lisa Kakinami, PhD, Concordia University; Eric Kennedy, PhD, York University; Sunmee Kim, PhD, University of Manitoba; Joanne Liu, PhD, McGill University; Colleen Norris, PhD, University of Alberta; Sandra Pelaez, PhD, Université de Montréal; Louise Pilote, MD, McGill University; Paul Poirier, MD, Université Laval; Justin Presseau, PhD, University of Ottawa; Eli Puterman, PhD, University of British Columbia; Joshua Rash, PhD, Memorial University; Paula AB Ribeiro, PhD, MBMC; Mohsen Sadatsafavi, PhD, University of British Columbia; Paramita Saha Chaudhuri, PhD, McGill University; Jovana Stojanovic, PhD, Concordia University; Eva Suarthana, MD, PhD, Université de Montréal / McGill University; Sze Man Tse, MD, CHU Sainte-Justine; Michael Vallis, PhD, Dalhousie University; CHILE: Nicolás Bronfman Caceres, PhD, Universidad Andrés Bello; Manuel Ortiz, PhD, Universidad de La Frontera; Paula Beatriz Repetto, PhD, Universidad Católica de Chile; COLOMBIA: Mariantonia Lemos-Hoyos, PhD, Universidad EAFIT; CYPRUS: Angelos Kassianos, PhD, University of Cyprus; DENMARK: Naja Hulvej Rod, PhD, University of Copenhagen; FRANCE: Mathieu Beraneck, PhD, Université de Paris; CNRS; Gregory Ninot, PhD, Université de Montpellier; GERMANY: Beate Ditzen, PhD, Heidelberg University; Thomas Kubiak, PhD, Mainz University; GHANA: Sam Codjoe MPhil,MSc, University of Ghana; Lily Kpobi, PhD, University of Ghana; Amos Laar, PhD, University of Ghana; GREECE: Theodora Skoura, PhD, Aretaieio Hospital Athens University; INDIA: Delfin Lovelina Francis, PhD, Vinayaka Mission’s Dental College; Naorem Kiranmala Devi, PhD, University of Delhi; Sanjenbam Meitei, PhD, Manipur University; Suzanne Tanya Nethan, MDS, School of Preventive Oncology; Lancelot Pinto, MD, PhD, Hinduja Hospital and Medical Research Centre; Kallur Nava Saraswathy, PhD, University of Delhi; Dheeraj Tumu, MD, World Health Organization (WHO); INDONESIA: Silviana Lestari, MD, PhD, Universitas Indonesia; Grace Wangge, MD, PhD, SEAMEO Regional Center for Food and Nutrition; IRELAND: Molly Byrne, PhD, National University of Ireland, Galway; Hannah Durand, PhD, National University of Ireland, Galway; Jennifer McSharry, PhD, National University of Ireland, Galway; Oonagh Meade, PhD, National University of Ireland, Galway; Gerry Molloy, PhD, National University of Ireland, Galway; Chris Noone, PhD, National University of Ireland, Galway; ISRAEL: Hagai Levine, MD, Hebrew University; Anat Zaidman-Zait, PhD, Tel-Aviv University; ITALY: Stefania Boccia, PhD, Università Cattolica del Sacro Cuore; Ilda Hoxhaj, MD, Università Cattolica del Sacro Cuore, Stefania Paduano, MSc, PhD, University of Modena and Reggio Emilia; Valeria Raparelli, PhD, Sapienza - University of Rome; Drieda Zaçe, MD, MSc, PhDc, Università Cattolica del Sacro Cuore; JORDAN: Ala’S Aburub, PhD, Isra University; KENYA: Daniel Akunga, PhD, Kenyatta University; Richard Ayah, PhD, University of Nairobi, School Public Health; Chris Barasa, MPH, University of Nairobi, School Public Health; Pamela Miloya Godia, PhD, University of Nairobi; Elizabeth W. Kimani-Murage, PhD, African Population and Health Research Center; Nicholas Mutuku, PhD, University of Kenya; Teresa Mwoma, PhD, Kenyatta University; Violet Naanyu, PhD, Moi University; Jackim Nyamari, PhD, Kenyatta University; Hildah Oburu, PhD, Kenyatta University; Joyce Olenja, PhD, University of Nairobi; Dismas Ongore, PhD, University of Nairobi; Abdhalah Ziraba, PhD, African Population and Health Research Center; MALAWI: Chiwoza Bandawe, PhD, University of Malawi; MALAYSIA: Loh Siew Yim, PhD, Faculty of Medicine, University of Malaya; NIGERIA: Ademola Ajuwon, PhD, University of Ibadan; PAKISTAN: Nisar Ahmed Shar, PhD, CoPI-National Center in Big Data & Cloud Computing; Bilal Ahmed Usmani, PhD, NED University of Engineering and Technology; PERU: Rosario Mercedes Bartolini Martínez, PhD, Instituto de Investigacion Nutricional; Hilary Creed-Kanashiro, M.Phil., Instituto de Investigacion Nutricional; PORTUGAL: Paula Simão, MD, S. Pneumologia de Matosinhos; RWANDA: Pierre Claver Rutayisire, PhD, University Rwanda; SAUDI ARABIA: Abu Zeeshan Bari, PhD, Taibah University; SLOVAKIA: Iveta Nagyova, PhD, PJ Safarik University - UPJS; SOUTH AFRICA: Jason Bantjes, PhD, University of Stellenbosch; Brendon Barnes, PhD, University of Johannesburg; Bronwyne Coetzee, PhD, University of Stellenbosch; Ashraf Khagee, PhD, University of Stellenbosch; Tebogo Mothiba, PhD, University of Limpopo; Rizwana Roomaney, PhD, University of Stellenbosch; Leslie Swartz, PhD University of Stellenbosch; SOUTH KOREA: Juhee Cho, PhD, Sungkyunkwan University; Man-gyeong Lee, PhDc, Sungkyunkwan University; SWEDEN: Anne Berman, PhD, Karolinska Institutet; Nouha Saleh Stattin, MD, Karolinska Institutet; SWITZERLAND: Susanne Fischer, PhD, University of Zurich; TAIWAN: Debbie Hu, MD, MSc, Tainan Municipal Hospital; TURKEY: Yasin Kara, MD, Kanuni Sultan Süleyman Training and Research Hospital, Istanbul; Ceprail Simsek, MD Health Science University; Bilge Üzmezoğlu, MD, University of Health Science; UGANDA: John Bosco Isunju, PhD, Makerere University School of Public Health; James Mugisha, PhD, University of Uganda; UK: Lucie Byrne-Davis, PhD, University of Manchester; Paula Griffiths, PhD, Loughborough University; Joanne Hart, PhD, University of Manchester; Will Johnson, PhD, Loughborough University; Susan Michie, PhD, University College London; Nicola Paine, PhD, Loughborough University; Emily Petherick, PhD, Loughborough University; Lauren Sherar, PhD, Loughborough University; USA: Robert M. Bilder, PhD, ABPP-CN, University of California, Los Angeles; Matthew Burg, PhD, Yale; Susan Czajkowski, PhD, NIH - National Cancer Institute; Ken Freedland, PhD, Washington University; Sherri Sheinfeld Gorin, PhD, University of Michigan; Alison Holman, PhD, University of California, Irvine; Jiyoung Lee, PhD, University of Alabama; Gilberto Lopez ScD, MA, MPH, Arizona State University and University of Rochester Medical Center; Sylvie Naar, PhD, Florida State University; Michele Okun, PhD, University of Colorado, Colorado Springs; Lynda Powell, PhD, Rush University; Sarah Pressman, PhD, University of California, Irvine; Tracey Revenson, PhD, University of New York City; John Ruiz, PhD, University of Arizona; Sudha Sivaram, PhD, NIH, Center for Global Health; Johannes Thrul, PhD, Johns Hopkins; Claudia Trudel-Fitzgerald, PhD, Harvard T.H. Chan School of Public Health; Abehaw Yohannes, PhD, Azusa Pacific University. **Students:** AUSTRALIA: Rhea Navani, BSc, Monash University; Kushnan Ranakombu, PhD, Monash University; BRAZIL: Daisuke Hayashi Neto, Unicamp; CANADA: Tair Ben-Porat, PhD, Tel Aviv University; Anda Dragomir, University of Quebec at Montreal (UQAM) and CIUSSS-NIM; Amandine Gagnon-Hébert, BA, UQAM; Claudia Gemme, MSc, UQAM; Vincent Gosselin Boucher, University of Quebec at Montreal (UQAM) and CIUSSS-NIM; Mahrukh Jamil, Concordia University and CIUSSS-NIM; Lisa Maria Käfer, McGill University; Ariany Marques Vieira, MSc, Concordia University; Tasfia Tasbih, Concordia University and CIUSSS-NIM; Robbie Woods, MSc, Concordia University; Reyhaneh Yousefi, Concordia University and CIUSSS-NIM; FRANCE: Tamila Roslyakova, Université de Montpellier; GERMANY: Lilli Priesterroth, Mainz University; ISRAEL: Shirly Edelstein, Hebrew University-Hadassah School of Public Health; Ruth Snir, Hebrew University-Hadassah School of Public Health; Yifat Uri, Hebrew University-Hadassah School of Public Health; NEW ZEALAND: Mohsen Alyami, University of Auckland; NIGERIA: Comfort Sanuade; SERBIA: Katarina Vojvodic, University of Belgrade. **Community Participants:** CANADA: Olivia Crescenzi; Kyle Warkentin; DENMARK: Katya Grinko; INDIA: Lalita Angne; Jigisha Jain; Nikita Mathur, Syncorp Clinical Research; Anagha Mithe; Sarah Nethan, Community Empowerment Lab.

## Supplement 1 - methods

### Component structure of the iCARE concerns module

To cluster COVID-19-related concerns, we performed a principal component analysis (PCA) on a polychoric correlation matrix of the 14 (Survey 1) and 18 (Survey 2) variables in the COVID-19 concerns module (ordinal scale). An orthogonal (varimax) rotation was done in order to distribute the factor loadings. We identified concern patterns based on the Kaiser criterion (eigenvalue>1.0), scree plot, and components interpretability.^11^ Items with loadings higher than 0.4 were used to interpret each component of COVID-19 concerns. We observed a three-factor structure that included: ‘**Health concerns**’ (items: *being infected myself; the impact of being infected on my health, including dying*; *infecting other people I live with; a person with whom I live with being infected; a family member with whom I do not share my home being infected; a friend with whom I do not share my home being infected; infecting other people in the community*), ‘**Personal financial concerns**’ (items: *losing my job / family income; losing my / family savings; not having enough money for food and/or rent; there not being enough food left on shelves for people to eat*), ‘**Social/economic concerns**’ (items: *being isolated from other people; my country going into an economic recession/depression; how long it will take for things to go back to normal*).

**Supplement 2 − Table S1.**
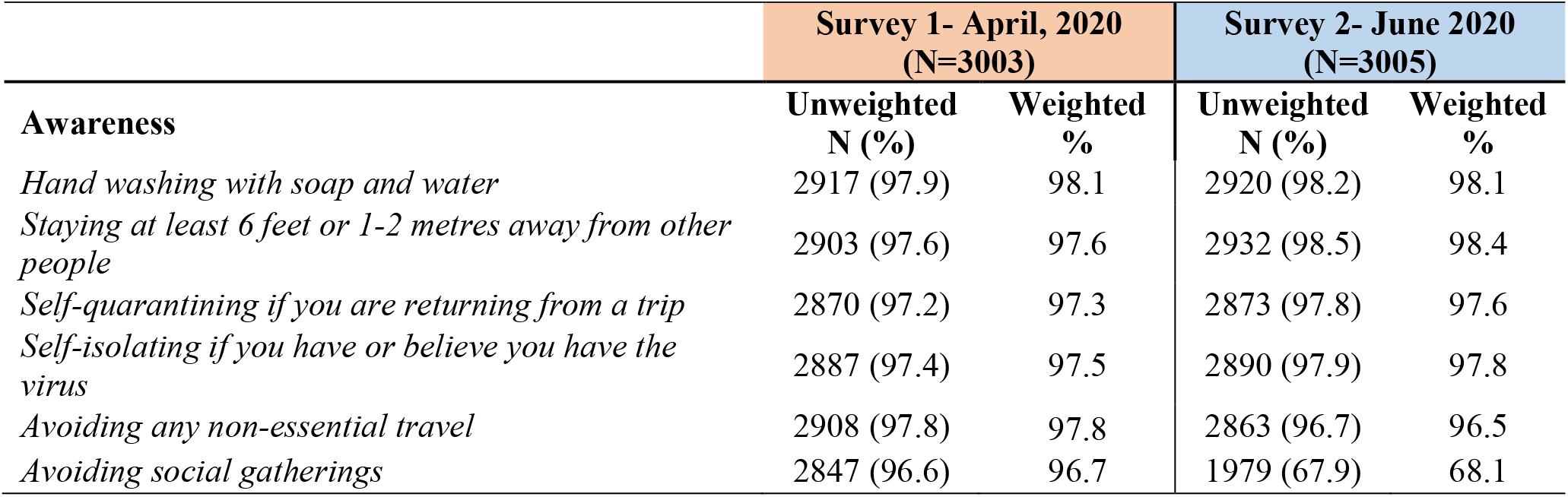
Overall frequency of awareness of the recommended preventive behaviours (presenting frequency and percentage of individuals that responded yes) by survey.

**Supplement 2 − Table S2.**
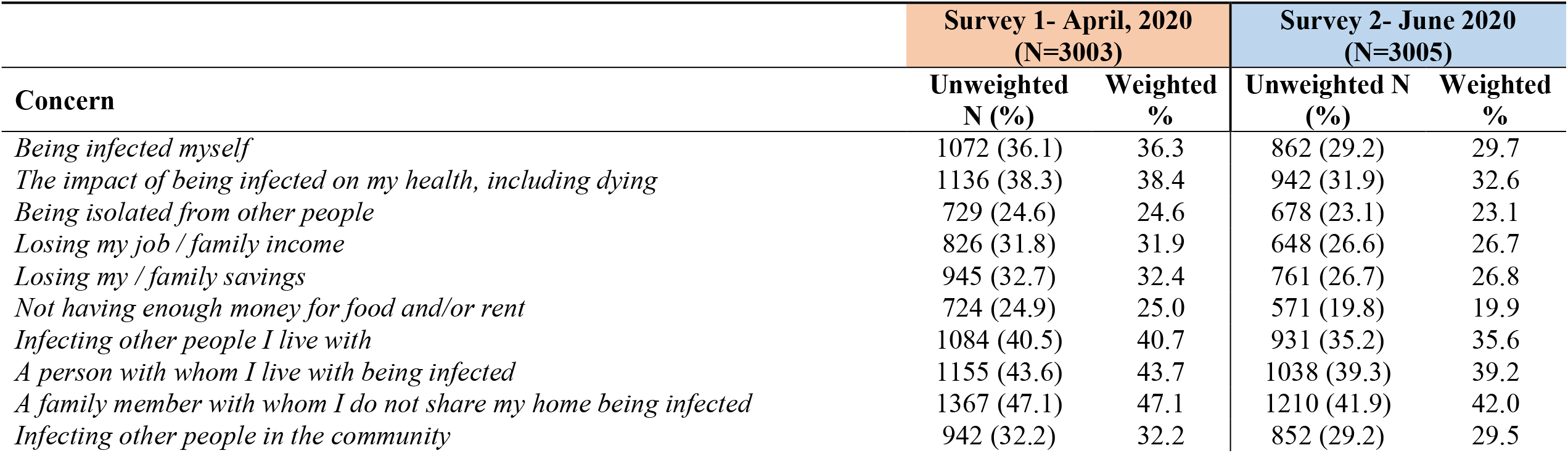
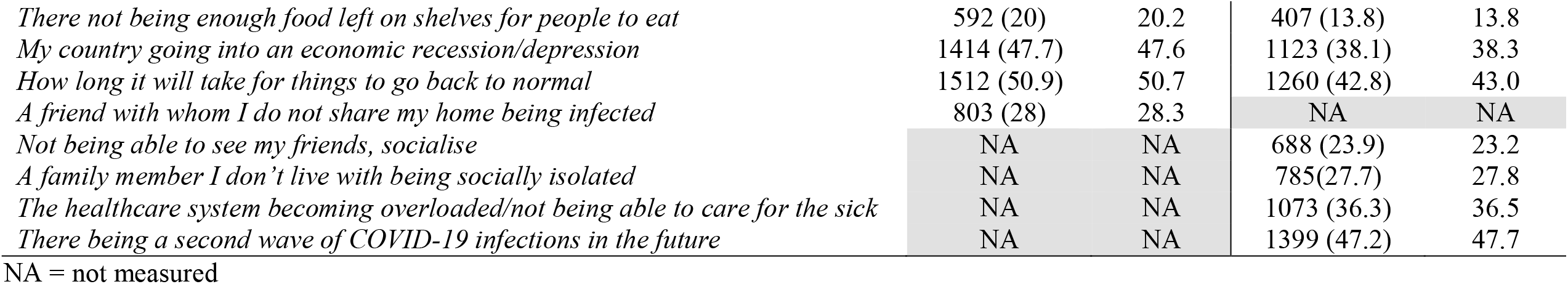
Overall frequency of COVID-19 related concerns (presenting frequency and percentage of individuals concerned *To a great extent*) by survey.

