## Supplemental Material for "Determinants of adherence to COVID-19 preventive behaviours in Canada: Results from the iCARE Study"

**Supplement 1 - methods**

### Component structure of the iCARE concerns module:

To cluster COVID-19-related concerns, we performed a principal component analysis (PCA) on a polychoric correlation matrix of the 14 (Survey 1) and 18 (Survey 2) variables in the COVID-19 concerns module (ordinal scale). An orthogonal (varimax) rotation was done in order to distribute the factor loadings. We identified concern patterns based on the Kaiser criterion (eigenvalue>1.0), scree plot, and components interpretability.[11] Items with loadings higher than 0.4 were used to interpret each component of COVID-19 concerns. We observed a three-factor structure that included: '**Health concerns**' (items: *being infected myself; the impact of being infected on my health, including dying*; *infecting other people I live with; a person with whom I live with being infected; a family member with whom I do not share my home being infected; a friend with whom I do not share my home being infected; infecting other people in the community*), '**Personal financial concerns**' (items: *losing my job / family income; losing my / family savings; not having enough money for food and/or rent; there not being enough food left on shelves for people to eat*), '**Social/economic concerns**' (items: *being isolated from other people; my country going into an economic recession/depression; how long it will take for things to go back to normal*).

**Supplement 2 – Table S1. Overall frequency of awareness of the recommended preventive behaviours (presenting frequency and percentage of individuals that responded yes) by survey**

| | Survey 1- April, 2020 (N=3003) | | Survey 2- June 2020 (N=3005) | |
|---|---|---|---|---|
| **Awareness** | **Unweighted N (%)** | **Weighted %** | **Unweighted N (%)** | **Weighted %** |
| *Hand washing with soap and water* | 2917 (97.9) | 98.1 | 2920 (98.2) | 98.1 |
| *Staying at least 6 feet or 1-2 metres away from other people* | 2903 (97.6) | 97.6 | 2932 (98.5) | 98.4 |
| *Self-quarantining if you are returning from a trip* | 2870 (97.2) | 97.3 | 2873 (97.8) | 97.6 |
| *Self-isolating if you have or believe you have the virus* | 2887 (97.4) | 97.5 | 2890 (97.9) | 97.8 |
| *Avoiding any non-essential travel* | 2908 (97.8) | 97.8 | 2863 (96.7) | 96.5 |
| *Avoiding social gatherings* | 2847 (96.6) | 96.7 | 1979 (67.9) | 68.1 |

**Supplement 2 – Table S2. Overall frequency of COVID-19 related concerns (presenting frequency and percentage of individuals concerned *To a great extent*) by survey**

| | Survey 1- April, 2020 (N=3003) | | Survey 2- June 2020 (N=3005) | |
|---|---|---|---|---|
| **Concern** | **Unweighted N (%)** | **Weighted %** | **Unweighted N (%)** | **Weighted %** |
| *Being infected myself* | 1072 (36.1) | 36.3 | 862 (29.2) | 29.7 |
| *The impact of being infected on my health, including dying* | 1136 (38.3) | 38.4 | 942 (31.9) | 32.6 |
| *Being isolated from other people* | 729 (24.6) | 24.6 | 678 (23.1) | 23.1 |
| *Losing my job / family income* | 826 (31.8) | 31.9 | 648 (26.6) | 26.7 |
| *Losing my / family savings* | 945 (32.7) | 32.4 | 761 (26.7) | 26.8 |
| *Not having enough money for food and/or rent* | 724 (24.9) | 25.0 | 571 (19.8) | 19.9 |
| *Infecting other people I live with* | 1084 (40.5) | 40.7 | 931 (35.2) | 35.6 |
| *A person with whom I live with being infected* | 1155 (43.6) | 43.7 | 1038 (39.3) | 39.2 |
| *A family member with whom I do not share my home being infected* | 1367 (47.1) | 47.1 | 1210 (41.9) | 42.0 |
| *Infecting other people in the community* | 942 (32.2) | 32.2 | 852 (29.2) | 29.5 |

| | | | | |
|---|---|---|---|---|
| *There not being enough food left on shelves for people to eat* | 592 (20) | 20.2 | 407 (13.8) | 13.8 |
| *My country going into an economic recession/depression* | 1414 (47.7) | 47.6 | 1123 (38.1) | 38.3 |
| *How long it will take for things to go back to normal* | 1512 (50.9) | 50.7 | 1260 (42.8) | 43.0 |
| *A friend with whom I do not share my home being infected* | 803 (28) | 28.3 | NA | NA |
| *Not being able to see my friends, socialise* | NA | NA | 688 (23.9) | 23.2 |
| *A family member I don't live with being socially isolated* | NA | NA | 785(27.7) | 27.8 |
| *The healthcare system becoming overloaded/not being able to care for the sick* | NA | NA | 1073 (36.3) | 36.5 |
| *There being a second wave of COVID-19 infections in the future* | NA | NA | 1399 (47.2) | 47.7 |

NA = not measured
